# Psychological distress of healthcare workers in Québec (Canada) during the second and the third pandemic waves

**DOI:** 10.1101/2021.10.19.21265175

**Authors:** Sara Carazo, Mariève Pelletier, Denis Talbot, Nathalie Jauvin, Gaston De Serres, Michel Vézina

**Affiliations:** CHU de Québec-Laval University Research Center, Québec, Québec, Canada; Biological and occupational risks unit. Quebec national institute of public health, Québec, Québec, Canada; Social and preventive medicine department, Faculty of Medicine, Laval University, Québec, Québec, Canada

**Keywords:** COVID-19, healthcare worker, psychological distress, K6 scale, workplace psychosocial risk, occupational risk

## Abstract

**Objective:** We aimed to measure the prevalence of psychological distress among Quebec healthcare workers (HCWs) during the second and third pandemic waves and to assess the effect of psychosocial risk factors (PSRs) on work-related psychological distress among SARS-CoV-2 infected (cases) and non-infected (controls) HCWs.

**Methods:** A self-administered survey was used to measure validated indicators of psychological distress (K6 scale) and PSR (questions based on Karasek and Siegrist models, value conflicts and work-life balance). Adjusted robust Poisson models were used to estimate prevalence ratios.

**Results:** 4068 cases and 4152 controls completed the survey. Prevalence of high work-related psychological distress was 42%; it was associated with PSRs (mainly work-life balance, value conflicts and high psychological demands) but not with SARS-CoV-2 infection.

**Conclusion:** Primary prevention measures targeting PSRs are needed to reduce mental health risks of HCWs.

## Introduction

Healthcare workers (HCWs) have been at high risk of exposure to SARS-CoV-2 since the beginning of the pandemic. In Québec, Canada, their risk of infection was 10 times higher than the general working age population during the first pandemic wave and 4 times higher during the second wave [1,2]. In addition, several studies have shown that beyond the risk of infection, the pandemic has also had a negative impact on the mental health of HCWs [3–10].

In the spring of 2020, Québec HCWs with COVID-19 reported high workload, feelings of abandonment (lack of support) or lack of recognition, which are psychosocial risk factors (PSR) known to impact on mental health at work [11]. Numerous studies have demonstrated the strong links between PSRs, such as high psychological demand, low support at work, low recognition, value conflicts and difficulty in balancing work and personal life, with psychological distress, an early indicator of mental disorders [11–16].

Given these initial findings, the current study was conducted during the second and third pandemic waves to estimate the prevalence of high and very high psychological distress, the proportion that is work-related, and to assess the effect of psychosocial risk factors on work-related psychological distress among HCWs infected or not with COVID-19.

## Methods

### Study population

This test-negative case-control study was conducted among HCWs tested for SARS-CoV-2 by PCR between November 15, 2020 and May 29, 2021. HCWs with a positive result were cases and controls were those with a negative result [2]. Inclusion criteria included: 1) being a HCW, defined as someone working in the health field or in a health facility; 2) having worked during the 14 days before symptom onset or testing; 3) aged 18 years or older; 4) able to communicate in French or English; 5) living in Québec. Eligible cases were identified from the provincial reportable disease database that includes all confirmed COVID-19 cases, while controls were identified from the provincial laboratory database which record all PCR tests done for SARS-CoV-2. These individuals were contacted by phone and invited to participate to the study between December 3, 2020 and July 31, 2021. Consenting participants had to complete a questionnaire which was mostly (98%) self-administered online or, for those not at ease with electronic questionnaire, completed during the call with the research assistant (2%).

### Data collection and measurement of psychosocial risk and psychological distress

Participants were asked about their socio-demographic characteristics (age, sex, race/ethnicity and household composition (only cases)), employment characteristics (occupation and type of facility) and infection prevention and control measures in their workplace.

In addition, PSRs were measured by validated indicators from the two main internationally recognized models, that is, Karasek and Theorell’s “demand-control-support” model and Siegrist’s “effort-reward imbalance” model [17,18]. The indicators associated with these two models are: the psychological demands (workload), decision authority (one of the two components of decision autonomy), job strain (combination of high psychological demands and low or moderate decision authority), support from coworkers and supervisors, and reward. The questions retained were those used in the Québec Population Health Survey 2014/15 (QPHS) in order to be able to compare the results [19]. Three questions were added to measure PSRs in the context of COVID-19. Two questions from surveys in France related to value conflicts assessed: 1) the perception of having the means to do quality work and, 2) the perception of having to work in a way that offends one’s professional conscience [12,14,20]. The third question was about work-life balance (see Supplemental Digital Content (SDC) - Annex 1 for questions and details regarding the indicators) [21].

Psychological distress during the 30 days prior to completing the questionnaire was measured with the Kessler (K6) scale [22]. It includes six questions scored between 0 and 4 for a maximum of 24 points. A score of ≥7 is considered indicative of high psychological distress: distress is high with a socre of 7 to 12 and very high with ≥13 (SDC Annex 1). Even though K6 is not a diagnostic tool, psychological distress is an early indicator of mental health illness, particularly of two of the most frequent syndromes: depression and anxiety [22–24]. Work-related psychological distress was identified through a question about the perceived link between the feelings reported and the current job.

### Statistical analyses

Prevalences and their 95% confidence intervals (CI) were estimated. Univariate associations between PSRs and COVID-19 status and work-related psychological distress were evaluated with a chi-square test.

To avoid convergence problems often encountered with log-binomial models [25–27], we used robust Poisson models, adjusted for sex, age, race/ethnicity, type of occupation and COVID-19 status, to estimate the prevalence ratios (PR) of high or very high psychological distress associated with the presence of occupational psychosocial risks. The independent association between each RPS, the COVID-19 status and psychological distress was measured in a model including all PSRs. The impact of the household composition was assessed in a model including only the cases, which showed no changes in the estimates. The association between the simultaneous presence of one to five PSRs and psychological distress was also examined with the construction of an overall indicator of RPS. Finally, an exploratory analysis evaluated the association of work-related psychological distress with the perceived risk of acquiring COVID-19 in the workplace (before contracting the disease) and some organization and infection prevention and control measures in the workplace.

### Ethical aspects

The study was conducted under the legal mandate entrusted to the National Institute of Public Health of Québec by the National Director of Public Health of Québec under the Public Health Act. The study was approved by the research ethics committee of the CHU (University Hospital Center) de Québec-Université Laval and all participants gave oral or written informed consent before inclusion.

## Results

### Study population

During the study period 23,318 SARS-CoV-2 laboratory confirmed infected HCWs were reported in Québec; 12,601 were successfully reached; 949 (7.5%) did not meet the inclusion criteria and 21.2% (2666) refused to participate. For controls, 11498 HCWs who tested negative for SARS-CoV-2 were reached among the 21,900 randomly selected from the laboratory database; 1243 (10.8%) were excluded and 2527 (22.0%) did not consent to participate. Of the 8986 cases and 7228 controls who agreed to participate, 4068 (45.3%) and 4152 (57.4%) respectively completed the survey as of 7 September 2021. The participation rate among eligible HCWs was therefore 34.9% for cases and 42.6% for controls. Cases were representative for age, sex and clinical characteristics of all infected HCWs reported in Québec during the study period [2].

From the 8220 participants, 83.3% were women, 66.7% were <45 years, 14.7% were born abroad, 85.2% defined themselves as white, 5.3% as black, 8.0% other ethnic/racial category and 1.5% did not answer the question. For occupation, 26.0% were nurses or nursing assistants, 19.1% patient healthcare support workers, 4.6% physicians and 11.6% worked in management or administration; 37.7% worked in acute-care hospitals, 16.8% in long-term care facilities (LTCF), 7.5% in private residences for elderly and 38.0% in other types of facilities.

Compared to controls, cases were more often men, older, defined themselves as black, and worked more often as patient healthcare assistants and in LTCFs (all p<0.01).

### Prevalence of psychological distress and psychosocial risks

High psychological distress (score ≥7) was reported by 50.7% of participant HCWs (53.1% of women and 39.5% of men), and 81.5% among them considered it as work-related. High work-related psychological distress was more frequent among controls (46.5%) than among SARS-CoV-2 infected HCWs (36.1%); and among nurses and nursing assistant (49.9%) than among other occupations (37.8% to 38.8%) (Figure 1). The monthly prevalence of high-work related psychological distress in the previous 30 days remained quite stable over the study period: between 29% and 38% among cases (Cochran-Armitage trend test p = 0.046) and between 44% and 49% among controls (trend test p = 0.16), with June and July 2021 being the months with lowest prevalences (Figure 2).

**Figure 1.**
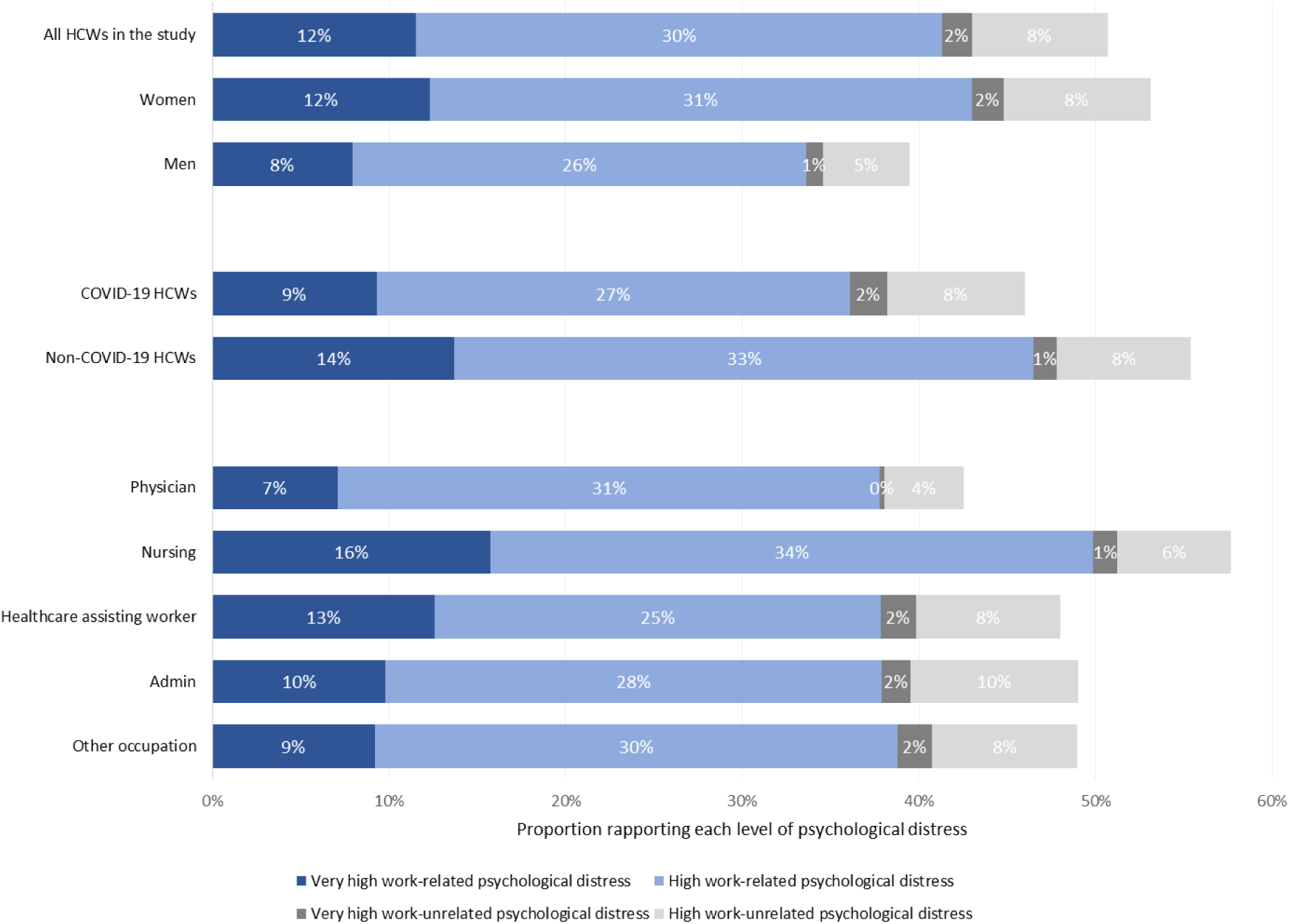
Level of psychological distress among SARS-CoV-2 infected healthcare workers as measured by the Kessler (K6) scale, stratified by sex, COVID-19 status and occupation Abbreviations: Admin= Administration and management staff; HCW= healthcare worker; Nursing= nurses and nursing assistants Note: High psychological distress = score of 7 to 12 in the K6 scale; Very high psychological distress = score 13 to 24 in the K6 scale

**Figure 2.**
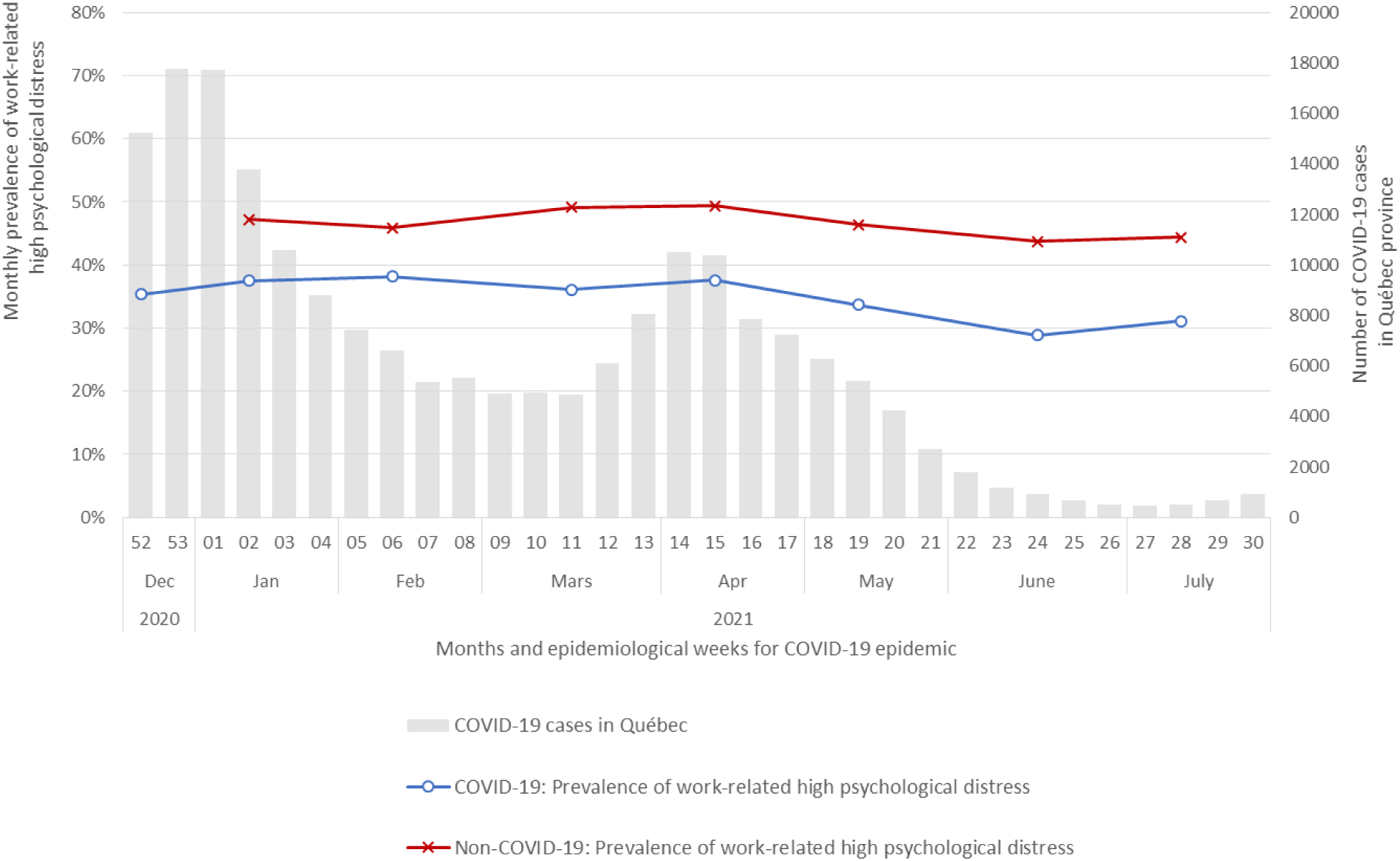
Monthly prevalence of work-related high psychological distress (K6 ≥7) among SARS-CoV-2 infected and non-infected healthcare workers by month of survey complexion, and weekly number of COVID-19 cases in Québec province

High psychosocial demands were reported by 38.6% (52.2% of nurses or nursing assistants), low or moderate decision authority by 52.1% (61.6% of healthcare support workers and 57.5% of nurses and nursing assistants), low reward by 33.8% (42.8% of healthcare support workers, 37.6% of nurses and nursing assistants but 4.7% of physicians), and low or moderate co-worker or supervisor support by 17.2% and 22.5% respectively. Overall, 77.5% reported not having (sometimes, often or always) the means to do quality work and this reached 87.2% among nurses and nursing assistants. Sometimes, often or always working against their professional conscience was reported by 52.2% of HCWs, and up to 69.9% among nurses and nursing assistants. Finally, 30.8% considered that they had difficulties to keep a work-life balance, a proportion reaching 47.5% among physicians (SDC - Tables S1, S2 and S3). Compared to controls, low or moderate decision authority was more often reported by cases (54.8% versus 49.5%), while difficult work-life balance and conflict values were less prevalent among cases (26.5%, 73.9% and 50.3% respectively versus 35.3%, 81.0% and 54.1% for controls) (SDC - Table S1).

### Association between psychosocial risks, COVID-19 status and work-related psychological distress

In univariate analysis, all PSRs were statistically associated with high and very high work-related psychological distress both among cases and among controls. High work-related psychological distress was more frequent among HCWs with difficult work-life balance (63.2% of cases and 71.0% of controls versus 26.5% and 33.0% respectively among those not exposed to this risk) and with high psychological demands (56.5% of cases and 67.1% of controls versus 24.0% and 32.8% respectively among those with low or moderate), followed by those with low reward, low or moderate supervisor support and work against their professional conscience. Very high work-related psychological distress was more prevalent (19% to 24%) among HCWs with difficult work-life balance, low or moderate co-worker or supervisor support, low reward and high psychological demands than among respondent not exposed to these risks (5% to 8%). Associations were similar for both sexes (SDC - Table S1).

When each psychosocial risk was analysed in a separate adjusted regression model including COVID-19 status, not having the means to do quality work had the strongest (2.5 times higher than those always having the means) association with high work-related psychological distress. Those working against their professional conscience (compared to never doing it), those with high psychological demands (compared to low or moderate) or difficult work-life balance (compared to easy) all had ≈2 times higher prevalence of psychological distress. The strength of association was greater (prevalence ratios between 2.1 and 4.3) when the outcome evaluated was very high work-related psychological distress. In sex-stratified analyses, prevalence ratios remained similar to global estimates. When stratified by COVID-19 status, the association between indicators of values conflict and psychological distress were stronger for cases than for controls (statistically significant only for working against one’s professional conscience) (Table 1).

**Table 1.**
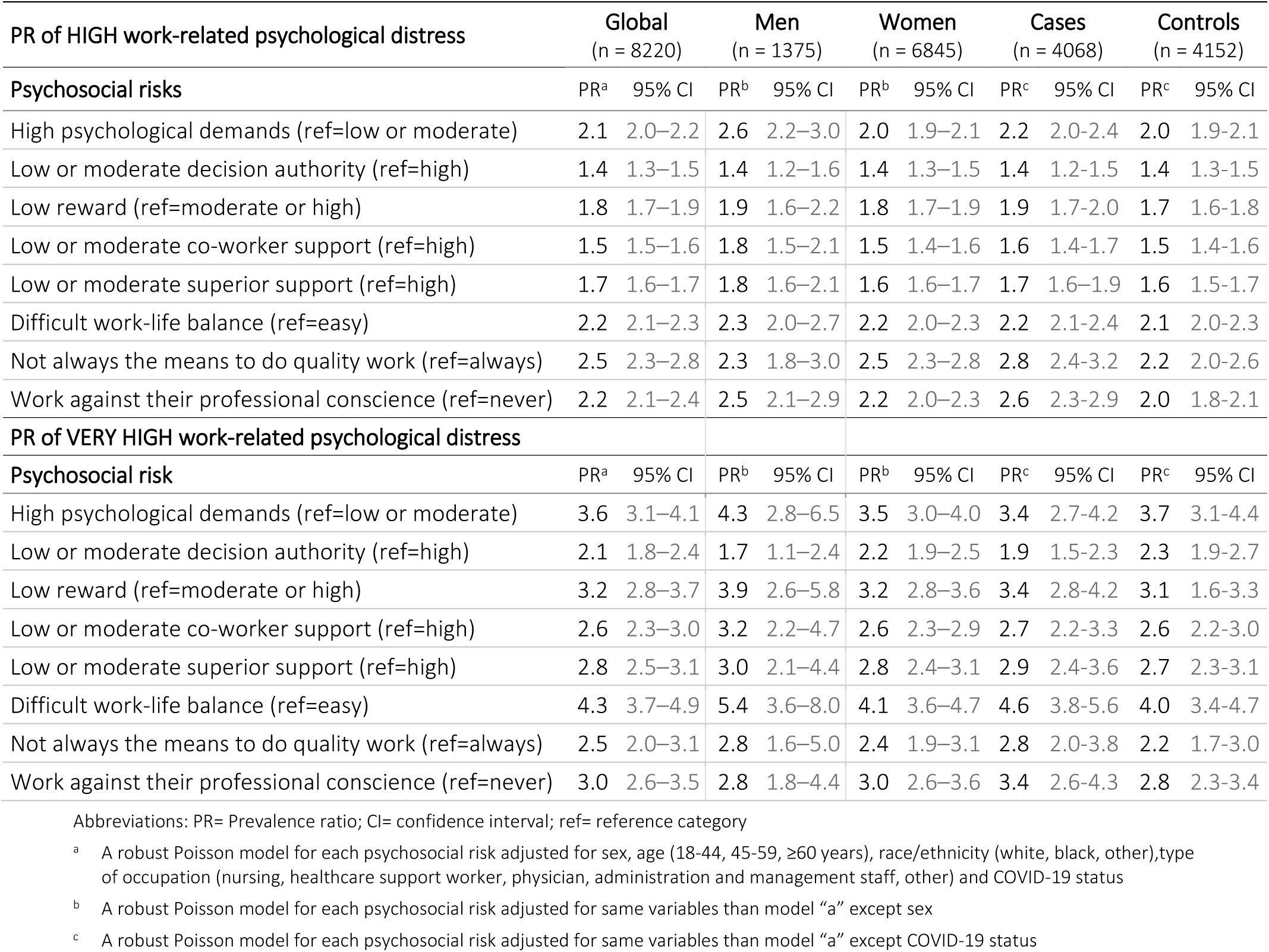
Prevalence ratios of high (K6 score ≥ 7) or very high (K6 score ≥ 13) work-related psychological distress according to each psychosocial risk, globally and stratified by sex (one model for each risk) and COVID-19 status

In the adjusted model that included all PSRs, associations were similar to those found in the separate models but prevalence ratios were lower and some lost statistical significance. The difficult work-life balance (PR=1.6, 95% CI: 1.5 – 1.7), the lack of means to do quality work (PR=1.6, 95% CI: 1.5 – 1.7), and working against one’s professional conscience (PR=1.5, 95% CI: 1.4 – 1.6) had the strongest associations with high work-related psychological distress, while the difficulty to have work-life balance was associated to the highest risk of very high work-related psychological distress (PR=2.7; 95% CI: 2.3 – 3.0). Having had COVID-19 was associate to a lower risk of high and very high psychological distress (Table 2).

**Table 2.** Prevalence ratios of high (K6 score ≥ 7) or very high (K6 ≥ 13) work-related psychological distress adjusted for all psychosocial risks and COVID-19 status, globally and stratified by sex

| PR of HIGH work-related psychological distress | Global<br>(n = 8220) |  | Men<br>(n = 1375) |  | Women<br>(n = 6845) |  |
| --- | --- | --- | --- | --- | --- | --- |
| Psychosocial risk | PR<sup>a</sup> | 95% CI | PR<sup>b</sup> | 95% CI | PR<sup>b</sup> | 95% CI |
| High psychological demands (ref=low or moderate) | 1.4 | 1.3 – 1.4 | 1.7 | 1.4 – 2.0 | 1.3 | 1.2 – 1.4 |
| Low or moderate decision authority (ref=high) | 1.0 | 1.0 – 1.1 | 1.0 | 0.8 – 1.1 | 1.0 | 1.0 – 1.1 |
| Low reward (ref=moderate or high) | 1.2 | 1.1 – 1.3 | 1.1 | 0.9 – 1.3 | 1.2 | 1.1 – 1.3 |
| Low or moderate co-worker support (ref=high) | 1.1 | 1.1 – 1.2 | 1.2 | 1.1 – 1.4 | 1.1 | 1.0 – 1.2 |
| Low or moderate superior support (ref=high) | 1.1 | 1.1 – 1.2 | 1.2 | 1.1 – 1.4 | 1.1 | 1.1 – 1.2 |
| Difficult work-life balance (ref=easy) | 1.6 | 1.5 – 1.7 | 1.7 | 1.5 – 2.0 | 1.6 | 1.5 – 1.7 |
| Not the means to do quality work (ref=always the means) | 1.6 | 1.5 – 1.8 | 1.4 | 1.1 – 1.8 | 1.7 | 1.5 – 1.9 |
| Work against their professional conscience (ref=never) | 1.5 | 1.4 – 1.6 | 1.7 | 1.4 – 2.0 | 1.5 | 1.4 – 1.6 |
| **COVID-19 status (ref=non-COVID-19)** | 0.9 | 0.8 – 0.9 | 0.9 | 0.8 – 1.0 | 0.9 | 0.8 – 0.9 |
| **PR of VERY HIGH work-related psychological distress** |  |  |  |  |  |  |
| Psychosocial risk | PR<sup>a</sup> | 95% CI | PR<sup>b</sup> | 95% CI | PR<sup>b</sup> | 95% CI |
| High psychological demands (ref=low or moderate) | 1.7 | 1.5 – 2.0 | 1.9 | 1.2 – 3.1 | 1.7 | 1.4 – 2.0 |
| Low or moderate decision authority (ref=high) | 1.2 | 1.1 – 1.4 | 0.9 | 0.7 – 1.3 | 1.3 | 1.1 – 1.5 |
| Low reward (ref=moderate or high) | 1.6 | 1.4 – 1.8 | 1.7 | 1.0 – 2.6 | 1.6 | 1.3 – 1.8 |
| Low or moderate co-worker support (ref=high) | 1.5 | 1.3 – 1.7 | 1.7 | 1.1 – 2.5 | 1.4 | 1.3 – 1.7 |
| Low or moderate superior support (ref=high) | 1.4 | 1.2 – 1.6 | 1.6 | 1.1 – 2.4 | 1.4 | 1.2 – 1.6 |
| Difficult work-life balance (ref=easy) | 2.7 | 2.3 – 3.0 | 3.5 | 2.3 – 5.2 | 2.6 | 2.2 – 3.0 |
| Not the means to do quality work (ref=always the means) | 1.1 | 0.9 – 1.4 | 1.1 | 0.6 – 2.0 | 1.1 | 0.9 – 1.4 |
| Work against their professional conscience (ref=never) | 1.6 | 1.4 – 1.9 | 1.4 | 0.9 – 2.1 | 1.6 | 1.4 – 1.9 |
| **COVID-19 status (ref=non-COVID-19)** | 0.8 | 0.7 – 0.9 | 0.9 | 0.6 – 1.3 | 0.7 | 0.7 – 0.8 |

Abbreviations: PR= Prevalence ratio; CI= confidence interval; ref= reference category
<sup>a</sup> A robust Poisson model including all psychosocial risks and COVID-19 status and adjusted for sex, age (18-44, 45-59, $\geq 60$ years), race/ethnicity (white, black, other) and type of occupation (nursing, healthcare support worker, physician, administration and management staff, other)
<sup>b</sup> A robust Poisson model including all psychosocial risks and COVID-19 status and adjusted for same variables than model “a” except sex

The exposure to one additional PSR was linearly associated with an increased prevalence of high and very high work-related psychological distress (only PSRs of Karasek’s and Siegrist’s models were considered). HCWs exposed to 4 or 5 PSRs had prevalence of high psychological distress 3.9 and 4.4 times greater than those not exposed to any PSR, respectively. Similarly, the prevalence of very high psychological distress was 18.0 and 28.2 times greater among HCWs exposed to 4 or 5 PSRs as compared to unexposed ones, respectively (Table 3).

**Table 3.**
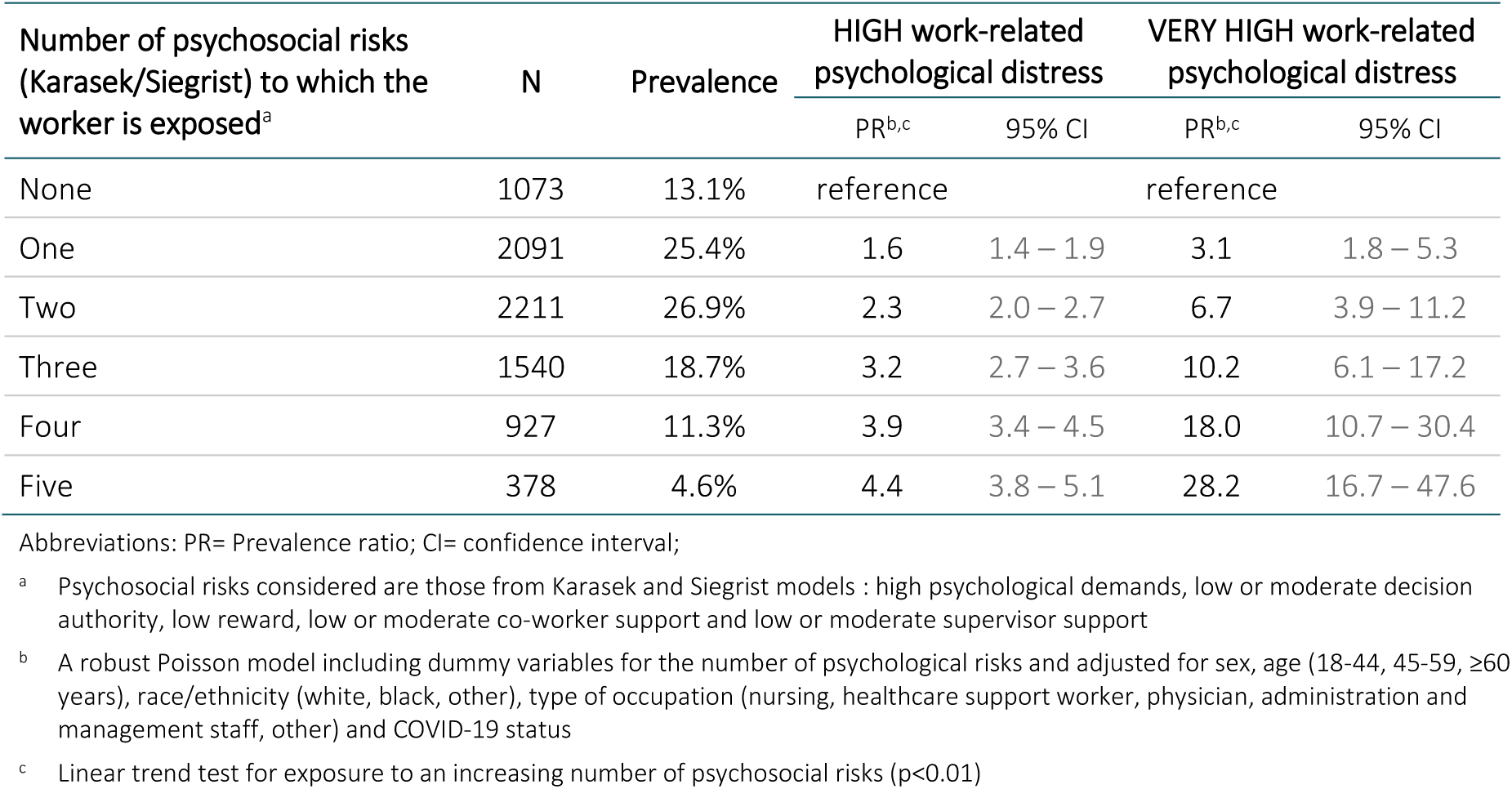
Prevalence ratios of high (K6 ≥ 7) or very high (K6 score ≥ 13) work-related psychological distress according to the global level of job psychosocial risk (number of risks)

High and very high work-related psychological distress was more common among workers who considered themselves to be at high or very high risk of acquiring COVID-19 in the workplace, as well as among those making a negative assessment of different infection prevention measures at work. When adjusted for demographic characteristics, perceived risk of acquiring COVID-19 in the workplace (PR=1.3; 95% CI: 1.2 – 1.4), lack of human resources (PR=1.5; 95% CI: 1.4 – 1.6), and persistence of COVID-19 symptoms for cases (PR=1.3; 95% CI: 1.2 – 1.4), were associated with high work-related psychological distress (SDC - Table S4).

## Discussion

Half (51%) of HCWs participating to the survey reported high or very high psychological distress, and 82% among them perceived their distress as work-related. Work-related high psychological distress was higher among non-infected HCWs (47%) compared to SARS-CoV-2 infected participants (36%). These rates remained quite stable over the study period, being 44% and 29% respectively at the end of the study period when COVID-19 incidence in the community was lowest. In addition, the results confirmed the original hypothesis that the prevalence of high and very high work-related psychological distress is associated with PSRs and not with SARS-CoV-2 infection, for both men and women. The PSRs most strongly associated with distress were: the difficulty balancing work and personal life, not having the means to do quality work, having to work in a way that offends one’s professional conscience, and high psychological demands. The prevalence of high and very high work-related psychological distress seemed to increase with the number of PSRs to which healthcare worker was exposed, with the prevalence of very high psychological distress being up to 28 times higher among HCW with five PSRs than among those with none.

A meta-analysis about the psychological impact of COVID-19 pandemic among HCWs reported a 12-study pooled prevalence of psychological distress of 46%, but the studies used different distress measurement tools, did not asses the relationship with work and were highly heterogeneous [3]. In our study, rates of work-related high and very high psychological distress among HCWs with COVID-19 working during the second and third pandemic waves were, respectively, more than 2 and 3 times higher than historical rates among workers of the same sector reported in the 2014/15 Québec population health survey using the same Kessler scale, questions and cut-off. These comparisons should be cautiously interpreted considering that the Québec population health survey data were collected 5 years earlier and outside the pandemic context. As described in other studies, nurses and women had higher prevalences of high and very high psychological distress than other job categories or men [3,28,29].

Our results are consistent with those of numerous prospective and cross-sectional epidemiological studies that have shown strong associations between exposure to almost all of the PSRs measured in our study and workers’ psychological distress, as well as depression, anxiety, and burnout [11,12,30–37].

This study underlines the importance of the association between high work-related psychological distress and not having the means to do a quality job and working in a way that harms professional integrity. These characteristic elements of value conflicts may be associated with moral distress or injury and may lead to a loss of professional identity and consequent loss of meaning in work [13,14]. Since the beginning of this health crisis, several authors have reported negative impact of exposure to highly demanding situations generating ethical or moral conflicts or dilemmas for caregivers: lack of resources to provide the care deemed necessary, disconnection of services with certain patients or users, conflicts between professional obligations and one’s own safety or that of one’s loved ones, difficult prioritization between the means at one’s disposal and the care to be provided, etc. [38–42].

These findings are concerning. Although psychological distress is not a mental illness, studies have shown that 80% of individuals with very high psychological distress scores also met DSM-4 diagnostic criteria for a mental disorder, such as anxiety or depression [23,24,43]. In addition, Pratt et al (2009) showed that very high psychological distress, as measured by a K6 questionnaire score of 13 and above, was associated with increased mortality of 30% compared to those with a score below 13 [44].

The importance of reducing the exposure of HCWs to PSRs, and more specifically to high psychological demands, is reinforced by the fact that the prevalence of very high work-related psychological distress increases with the number of PSRs to which workers were exposed. This observation is in line with theoretical models of PSRs, according to which the combination of high psychological demands, low autonomy and low social support at work represents a higher risk of health damage than exposure to just one of these three PSRs [17]. The same is true for the combination of high psychological demands and a low level of recognition at work [18].

The interpretations regarding primary prevention perspectives should be taken with caution because the cross-sectional, observational design of this study, does not allow to conclude on a causal relationship between PSRs and work-related psychological distress. However, our results suggest that the risk of work-related psychological distress could be reduced with the diminution of PSRs to which HCWs are exposed. For example, the risk of very high psychological distress may be reduced when work schedules are considered to facilitate work-life balance. Such results make it possible to consider action targets that could reduce the risk of mental health problems among healthcare workers. Indeed, during a pandemic, it may be more difficult to reduce the level of psychological work demands, a factor particularly associated with high and very high work-related psychological distress. However, the implementation of measures to increase the social support from the manager for his or her team or to recognize the efforts of workers could help reduce the rates of high and very high work-related psychological distress. These results are consistent with those of another study which showed that the prevalence of psychological distress and high depressive symptoms related to work are halved when exposure to emotionally demanding work is accompanied by a good level of decision latitude and social support at work [45]. According to another study, it would be possible to eliminate 14% of new cases of common mental disorders by reducing stressful situations at work [46]. Nevertheless, the association between psychological work demands and work-related distress remains highly significant, and actions that directly address this risk factor will have a greater likelihood of reducing psychological distress.

Work-related psychological distress among HCWs is important to understand the reasons for turnover and attrition in the health sector. A large American survey showed that, even before the pandemic, mental health problems such as burnout were cited by more than 30% of nurses who left their jobs, and that those who worked more than 40 hours per week were three times more likely to cite burnout as a reason for leaving their jobs. In addition, about two-thirds of respondents who had left or who considered leaving their jobs because of burnout attributed the causes to a stressful work environment or understaffing [47]. A Québec report published on September 2021 by the state commissioners of the nursing profession, concluded that to be able to offer quality care to the population, nurses must benefit from working conditions that respect their health, safety and integrity. Long working hours, compulsory overtime has to be abandoned, but the report also indicates that to promote the retention of nurses, employers must give them more autonomy to influence the management of care, as well as conditions that enhance their profession [48]. The impact of PSRs on staff sickness absence is also an issue raised in many studies. In a systematic review, Duchaine et al (2021) reported 14 prospective studies, in which several thousand workers were followed between one and twelve years, that demonstrated the impact of PSRs on certified work absences for a mental health problem [49].

One of the main strengths of the current study is its large number of participants, representative of all SARS-CoV-2 infected HCWs of Québec during the study period. Moreover, the survey took place between December 2020 and July 2021, which means that the results presented reflect the reality during the second and third pandemic waves in Québec and not a one-off situation. This study uses an original strategy in the choice of certain indicators that had not been used in other Québec or international occupational health surveys. These indicators made it possible to highlight relevant results that can broaden the perspectives for action to prevent psychological distress in the workplace, as value conflict issues.

This study has also some limitations. As said earlier, an observational cross-sectional survey cannot conclude on a causal relationship between PSRs and work-related psychological distress, but our results are consistent with several published studies. Exposures and events were self-reported, but bias should be limited because the questions were derived from theoretical models that have been internationally recognized for many years and used in national surveys. We cannot rule out a selection bias related to the ∼40% response rate, but such a bias would most likely underestimate the rate of distress as individuals with high and very high psychological distress may feel too bad to respond to a questionnaire.

## Conclusion

The psychological distress of health workers during the second and the third pandemic waves in Québec was mainly work-related, was not increased by having been infected by SARS-CoV-2, but was mostly associated with factors related to workload, such as high psychological demands, difficulties in balancing work and personal life, the lack of means to work in accordance with one’s professional conscience, and the lack of resources to ensure the quality of services to patients and to protect the safety of workers. Primary prevention measures targeting PSRs may reduce the psychological distress and mental health risks of these essential workers.

## Supporting information

Annex

## Data Availability

All data produced in the present work are contained in the manuscript

## List of Supplemental Digital Content

### Supplementary Digital Content – Tables.doc

**Table S1**. Prevalence of psychosocial risks (PSR) among SARS-CoV-2 infected and non-infected healthcare workers and association with work-related psychological distress

**Table S2**. Prevalence of psychosocial risks and items asked to measure them

**Table S3**. Prevalence of psychosocial risks among SARS-CoV-2 infected healthcare workers by sex and type of occupation and comparison with 2014/15 Québec’s population health survey (QPHS)

**Table S4**. Prevalence ratios of high (Kessler scale score ≥ 7) work-related psychological distress according to the perception of the risk of acquiring COVID-19 at work and of some infection prevention and control measures

### Supplementary Digital Content – Annex.doc

**Annex 1**. Questions in the survey for the items and variables included in the study and construction of indicators (psychosocial risks of work and psychological distress)

## Notes

### Competing Interest Statement

DT is supported by a career award from the Fonds de recherche du Quebec - Sante. GDS received a grant from Pfizer for anti-meningococcal immunogenicity study not related to this study and reports funding by the Ministry of Health of Quebec to the Public Health Institute to develop the research project (paid to institution). The rest of the authors have nothing to declare

### Funding Statement

This work was supported by the Ministry of Health and Social Services of Quebec

### Author Declarations

The study was approved by the research ethics committee of the CHU (University Hospital Center) de Quebec-Universite Laval

