## Supplementary material for "Psychological distress of healthcare workers in Québec (Canada) during the second and the third pandemic waves": Annex

**Supplementary Digital Content - Annex**. Questions in the survey for the items and variables included in the study and construction of indicators (psychosocial risks of work and psychological distress)

| Items and Questions | Choice of answers and score assigned to each answer | Construction of indicators |
|---|---|---|
| **1. Psychosocial risks of work** | | |
| *The questions below relate to your current MAIN employment and the relationships with your professional environment.* | | |
| *A. I'm asked to do an excessive amount of work.* | Strongly disagree (0); Disagree (0); Agree (1); Strongly agree (2) | **Psychological demands.**<br>Sum of responses to items A to E (scale from 0 to 10)<br>Low level: score = 0 or 1<br>Moderate: score = 2 or 3<br>High level: score = 4 or more |
| *B. I receive contradictory requests (opposite) from others; these requests can come from different groups: supervisors, colleagues, customers, etc.* | Strongly disagree (0); Disagree (0); Agree (1); Strongly agree (2) | |
| *C. My job requires me to work very fast.* | Strongly disagree (0); Disagree (0); Agree (1); Strongly agree (2) | |
| *D. I have enough time to do my work.* | Strongly disagree (0); Disagree (0); Agree (1); Strongly agree (2) | |
| *E. My job requires me to work very hard (mental or physical requirements).* | Strongly disagree (0); Disagree (0); Agree (1); Strongly agree (2) | |
| *F. I experience many interruptions and distractions while performing my tasks.* | Strongly disagree (0); Disagree (0); Agree (1); Strongly agree (2) | |
| *G. I have the freedom to decide how I do my job.* | Strongly disagree (0); Disagree (0); Agree (1); Strongly agree (2) | **Decision-making authority**.<br>Sum of responses to items G and H (scale from 0 to 2)<br>High level: score = 2<br>Low or moderate level: score 0 or 1 |
| *H. I can influence how things are done at my job.* | Strongly disagree (0); Disagree (0); Agree (1); Strongly agree (2) | |
| *I. My job security is poor.* | Strongly disagree (0); Disagree (0); Agree (1); Strongly agree (2) | **Reward.**<br>Sum of responses to items I to L (scale from 0 to 8)<br>High level: score = 0<br>Moderate: score = 1 or 2<br>Low level: score = 3 or more |
| *J. Given all my efforts and accomplishments, I receive the respect and esteem that I deserve at my job.* | Strongly disagree (0); Disagree (0); Agree (1); Strongly agree (2) | |
| *K. Given all my efforts and accomplishments, my promotion prospects are satisfactory.* | Strongly disagree (0); Disagree (0); Agree (1); Strongly agree (2) | |
| *L. Given all my efforts and accomplishments, my salary is satisfactory.* | Strongly disagree (0); Disagree (0); Agree (1); Strongly agree (2) | |
| *M. My colleagues facilitate the execution of my work.* | Strongly disagree (0); Disagree (0); Agree (1); Strongly agree (2) | **Support from the colleagues.**<br>Sum of responses to items M and N (scale from 0 to 2)<br>High level: score = 2<br>Low or moderate level: score 0 or 1 |
| *N. At my job, I feel like I'm part of a team.* | Strongly disagree (0); Disagree (0); Agree (1); Strongly agree (2) | |
| *O. My immediate supervisor facilitates the execution of my work.* | Strongly disagree (0); Disagree (0); Agree (1); Strongly agree (2) | **Support from the supervisor.**<br>Sum of the responses to items O and P (scale from 0 to 2)<br>High level: score = 2<br>Low or moderate level: score 0 or 1 |
| *P. My immediate supervisor pays attention to what I say.* | Strongly disagree (0); Disagree (0); Agree (1); Strongly agree (2) | |
| *The questions below relate to your assessment of your current MAIN employment:* | | |

| Items and Questions | Choice of answers and score assigned to each answer | Construction of indicators |
|---|---|---|
| *A. I have the means to do a quality job.* | Never; Sometimes; Often; Always | **Not having the means to do quality work.** Yes: Answers never, sometimes or often |
| *B. I was forced to work in a way that offended my professional conscience.* | Never; Sometimes; Often; Always | **Forced to work in ways that offend their professional conscience.** Yes: Answers sometimes, often or always |
| **C. Work-Life Balance:** *How easy or difficult is it for you to maintain a balance between your work obligations and your personal or family responsibilities?* | Very easy; Easy; Neither easy nor difficult; Difficult; Very difficult; Don't know; Don't answer | **Difficulty balancing work and personal life.** Yes: Difficult or very difficult answers to the question |
| **2. Psychological distress** | | |
| *Over the last month, how often did you feel...* | | |
| *A. nervous?* | Never; Rarely; Sometimes; Most of the time; All the time; Don't know; Don't answer | **Psychological distress.** Sum of responses provided to questions A through F (0-4 points per question).<br>High distress = score ≥ 7<br>Very high distress = score ≥ 13 |
| *B. desperate?* | Never; Rarely; Sometimes; Most of the time; All the time; Don't know; Don't answer | |
| *C. agitated or unable to stay put?* | Never; Rarely; Sometimes; Most of the time; All the time; Don't know; Don't answer | |
| *D. so depressed that nothing could make you smile?* | Never; Rarely; Sometimes; Most of the time; All the time; Don't know; Don't answer | |
| *E. that everything was an effort (so much tired that everything requires an effort)?* | Never; Rarely; Sometimes; Most of the time; All the time; Don't know; Don't answer | |
| *F. worthless?* | Never; Rarely; Sometimes; Most of the time; All the time; Don't know; Don't answer | |
| Link with the work: *Do you believe that the feelings of last month are related to your current (main) employment?* | Completely related to my current (main) employment; Partially related to my current (main) employment most of the time; No at all related to my current (main) employment; Don't know; Don't answer | **Work-related distress:** if the participant answers completely or partially related to my current job. |
| **3. Sociodemographic and employment variables** | | |
| *Age* | Number of years | |
| *Gender* | M; F | |
| Ethnicity: *Which of the following category describes you best?* | Aboriginal (First Nations, Inuit, Métis); Caucasian; Asian; Black; Arab; Hispanic; Other category; Don't know; Prefer not to answer | |
| Family composition: *Other than you, how many adults (18 years old or older) live with you??*<br>*How many children (less than 18 years old) live with you (including the children in shared custody)?* | Number of adults and number of children | |
| Type of employment: *What is the main position that you hold in the healthcare system?* | Drop-down menu | |

| Items and Questions | Choice of answers and score assigned to each answer | Construction of indicators |
|---|---|---|
| Facility: *In what kind of facility did you mostly work during the 14 days before the onset of your illness?* | Drop-down menu | |
| **4. Perceptions of the risk factors and protection factors in the workplace** | | |
| Risk of COVID-19: *Since July 12, 2021, before getting sick, what was, in your opinion, your risk of catching COVID-19 in your workplace?* | Low; Moderate; High; Very high. | |
| *How would you assess the following factors in terms of COVID-19 prevention in your workplace at the moment?* | | |
| *A. The personal protection equipment (mask, eye protection/visor, gown, gloves) is available and easy to access.* | Strongly disagree; Disagree; Agree; Strongly agree | Agree = Agree or Strongly agree<br>Disagree = Disagree or Strongly disagree |
| *B. I have good training in COVID-19 prevention, which allows me to protect myself well.* | Strongly disagree; Disagree; Agree; Strongly agree | |
| *C. My training included practical exercises on using personal protection equipment* | Strongly disagree; Disagree; Agree; Strongly agree | |
| *D. I easily get the information that allows me to know who are the patients with COVID-19 or suspected to have COVID-19.* | Strongly disagree; Disagree; Agree; Strongly agree | |
| *E. The access to COVID-19 screening tests and to their result is easy for healthcare workers.* | Strongly disagree; Disagree; Agree; Strongly agree | |
| *F. The location and the layout of the furniture allow for a distancing of two meters with the other workers during breaks and meals.* | Strongly disagree; Disagree; Agree; Strongly agree | |
| *G. The human resources are sufficient to provide the patient care and ensure the safety of the workers* | Strongly disagree; Disagree; Agree; Strongly agree | |
