## Supplementary tables for "Psychological distress of healthcare workers in Québec (Canada) during the second and the third pandemic waves"

**Supplementary Digital Content -**

**Table S1.** Prevalence of psychosocial risks (PSR) among SARS-CoV-2 infected and non-infected healthcare workers and association with work-related psychological distress

| | COVID-19 HCWs | | | | Non-COVID-19 HCWs | | | |
|---|---|---|---|---|---|---|---|---|
| | Prevalence of PSR | High work-related psychological distress | | | Prevalence of PSR | High work-related psychological distress | | |
| | Total | Total | Men | Women | Total | Total | Men | Women |
| N | 4068 | 4068 | 838 | 3230 | 4152 | 4152 | 537 | 3615 |
| Psychosocial risks | Column % | Line % | Line % | Line % | Column % | Line % | Line % | Line % |
| High psychological demands | 37.5 | 56.5 | 53.4 | 57.2 | 39.8 | 67.1 | 61.2 | 68.0 |
| *Low or moderate psychological demands* | *62.5* | *24.0* | *17.1* | *25.9* | *60.2* | *32.8* | *25.6* | *33.9* |
| Low or moderate decision authority | 54.8 | 41.3 | 34.1 | 42.9 | 49.5 | 54.6 | 44.4 | 56.0 |
| *High decision authority* | *45.2* | *30.0* | *25.7* | *31.3* | *50.6* | *38.5* | *34.9* | *39.0* |
| Low reward | 32.5 | 53.7 | 48.2 | 55.0 | 35.0 | 63.7 | 53.1 | 65.1 |
| *Moderate or high reward* | *67.5* | *27.7* | *22.0* | *29.4* | *65.0* | *37.2* | *32.9* | *37.8* |
| Low or moderate co-worker support | 16.9 | 51.1 | 51.2 | 51.1 | 17.4 | 63.6 | 56.8 | 64.6 |
| *High co-worker support* | *83.1* | *33.1* | *26.2* | *35.0* | *82.6* | *42.8* | *35.8* | *43.9* |
| Low or moderate supervisor support | 22.6 | 55.5 | 50.3 | 56.6 | 22.5 | 66.4 | 60.3 | 67.4 |
| *High supervisor support* | *77.4* | *30.5* | *24.9* | *32.0* | *77.6* | *40.7* | *33.1* | *41.8* |
| Difficult work-life balance | 26.3 | 63.2 | 53.0 | 65.8 | 35.3 | 71.0 | 64.0 | 72.0 |
| *Easy work-life balance* | *73.8* | *26.5* | *21.8* | *27.8* | *64.7* | *33.0* | *28.0* | *33.8* |
| Not the means to do quality work | 73.9 | 43.9 | 36.5 | 45.8 | 81.0 | 52.1 | 44.7 | 53.1 |
| *Having the means to do quality work* | *26.1* | *14.2* | *11.6* | *14.9* | *19.0* | *22.3* | *22.4* | *22.3* |
| Work against professional conscience | 50.3 | 53.1 | 46.9 | 54.6 | 54.1 | 60.5 | 54.4 | 61.3 |
| *Not working against professional conscience* | *49.7* | *19.0* | *15.1* | *20.2* | *45.9* | *29.9* | *24.5* | *30.8* |
| | Prevalence of PSR | Very high work-related psychological distress | | | Prevalence of PSR | Very high work-related psychological distress | | |
| | Total | Total | Men | Women | Total | Total | Men | Women |
| Psychosocial risks | Column % | Line % | Line % | Line % | Column % | Line % | Line % | Line % |
| High psychological demands | 37.5 | *17.1* | *16.0* | *17.4* | 39.8 | *24.8* | *15.8* | *26.1* |
| *Low or moderate psychological demands* | *62.5* | *4.7* | *2.8* | *5.2* | *60.2* | *6.4* | *4.3* | *6.7* |
| Low or moderate decision authority | 54.8 | *12.0* | *9.4* | *12.6* | 49.5 | *19.3* | *11.2* | *20.5* |
| *High decision authority* | *45.2* | *6.1* | *5.5* | *6.3* | *50.6* | *8.2* | *6.5* | *8.4* |
| Low reward | 32.5 | *18.3* | *17.1* | *18.6* | 35.0 | *24.8* | *15.4* | *26.1* |
| *Moderate or high reward* | *67.5* | *5.0* | *3.2* | *5.5* | *65.0* | *7.7* | *5.5* | *8.0* |
| Low or moderate co-worker support | 16.9 | *19.7* | *17.9* | *20.0* | 17.4 | *27.5* | *20.0* | *28.7* |
| *High co-worker support* | *83.1* | *7.3* | *5.6* | *7.7* | *82.6* | *10.8* | *6.3* | *11.4* |
| Low or moderate supervisor support | 22.6 | *19.9* | 14.7 | 21.0 | 22.5 | *27.0* | *20.6* | *28.0* |

| | | | | | | | | |
|---|---|---|---|---|---|---|---|---|
| *High supervisor support* | *77.4* | *6.3* | *5.6* | *6.4* | *77.6* | *9.8* | *5.1* | *10.5* |
| Difficult work-life balance | 26.3 | *22.4* | 20.0 | 23.0 | 35.3 | *26.6* | *18.6* | *27.7* |
| *Easy work-life balance* | *73.8* | *4.7* | *3.1* | *5.1* | *64.7* | *6.6* | *4.1* | *7.0* |
| Not the means to do quality work | 73.9 | *11.3* | 9.1 | 11.9 | 81.0 | *15.4* | *10.0* | *16.1* |
| *Having the means to do quality work* | *26.1* | *3.8* | *2.7* | *4.1* | *19.0* | *6.5* | *4.8* | *6.8* |
| Work against professional conscience | 50.3 | *14.6* | 12.1 | 15.2 | 54.1 | *19.6* | *12.5* | *20.6* |
| *Not working against professional conscience* | *49.7* | 4.0 | *3.3* | *4.2* | *45.9* | *6.7* | *4.9* | *7.0* |

Abbreviations: PSR=Psychosocial risk

Note 1: The psychosocial risks identified refer to Karasek's job demand-control-support model and Siegrist's effort-reward imbalance model, as well as to conflicts of values and difficulties in reconciling work and family.

Note 2: Chi-square test of bivariate association between each psychosocial risk and work-related psychological distress <0.05 for all the proportions in the table; chi-square test of bivariate association between each psychosocial risk and COVID-19 status <0.05 for all proportions except co-worker and supervisor support.

**Table S2**. Prevalence of psychosocial risks and items asked to measure them

| Psychosocial risks and items to measure them | Men<br>n = 1375<br>% | Women<br>n = 6845<br>% | Total<br>n = 8220<br>% |
|---|---|---|---|
| **1. Level of psychological demands** | | | |
| Low | 33.4 | 31.2 | 31.6 |
| Moderate | 30.0 | 29.8 | 29.8 |
| High | 36.6 | 39.1 | 38.6 |
| **Items to measure the level of psychological demands** | | | |
| 1.1 Constant pressure due to a heavy workload | 37.0 | 40.9 | 40.2 |
| 1.2 Conflicting demands | 32.7 | 32.1 | 32.2 |
| 1.3 Work that requires working very quickly | 56.5 | 56.0 | 56.1 |
| 1.4 Not enough time to do the job | 32.6 | 38.5 | 37.5 |
| 1.5 Work that requires hard work (mental of physical demands) | 74.6 | 76.0 | 75.7 |
| **2. Level of decision authority (or job control)** | | | |
| Low or moderate | 49.2 | 52.7 | 52.1 |
| High | 50.8 | 47.3 | 47.9 |
| **Items to measure the level of decision authority** | | | |
| 2.1 Lack of choice over the way the job is done | 67.6 | 65.5 | 65.8 |
| 2.2 Lack of influence in decisions about his/her work | 64.3 | 60.7 | 61.3 |
| **3. Job strain** | | | |
| Yes | 21.8 | 24.8 | 24.3 |
| No | 78.2 | 75.2 | 75.7 |
| **4. Level of reward** | | | |
| Low | 31.0 | 34.3 | 33.8 |
| Moderate | 37.7 | 39.7 | 39.4 |
| High | 31.4 | 26.0 | 26.9 |
| **Items to measure the level of reward** | | | |
| 4.1 Job insecurity | 17.8 | 18.0 | 18.0 |
| 4.2 Poor job promotion prospects | 45.1 | 49.9 | 49.1 |
| 4.3 Unsatisfactory salary/income | 50.4 | 55.8 | 54.9 |

| Psychosocial risks and items to measure them | Men<br>n = 1375<br>% | Women<br>n = 6845<br>% | Total<br>n = 8220<br>% |
|---|---|---|---|
| 4.4 Do not receive the respect and prestige deserved at work | 25.2 | 25.9 | 25.7 |
| **5. Level of co-worker support** | | | |
| Low or moderate | 15.9 | 17.4 | 17.2 |
| High | 84.2 | 82.6 | 82.9 |
| **Items to measure the level of coworkers support** | | | |
| 5.1 No or little help and support from coworkers | 9.5 | 12.2 | 11.8 |
| 5.2 Little or no feeling of being part of a team at work | 10.9 | 10.5 | 10.6 |
| **6. Level of supervisor support** | | | |
| Low or moderate | 21.0 | 22.9 | 22.5 |
| High | 79.0 | 77.2 | 77.5 |
| **Items to measure the level of supervisor support** | | | |
| 6.1 No or little help and support from the immediate superior | 16.2 | 18.1 | 17.8 |
| 6.2 Immediate superior is rarely or never willing to listen | 17.0 | 18.1 | 17.9 |
| **7. Work and family/personal life conflict** (difficulty maintaining a balance between work obligations and personal and family responsibilities) | | | |
| Yes | 28.2 | 31.3 | 30.8 |
| No | 71.9 | 68.7 | 69.2 |
| **8. Do not have the means to do quality work** (always, often or sometimes) | 74.6 | 78.1 | 77.5 |
| **9. Forced to work in a way that offends their professional conscience** (always, often or sometimes) | 48.0 | 53.1 | 52.2 |

Note 1: The psychosocial risks identified refer to Karasek's job demand-control-support model and Siegrist's effort-reward imbalance model, as well as to conflicts of values and difficulties in balancing work and family.

**Table S3**. Prevalence of psychosocial risks among SARS-CoV-2 infected healthcare workers by sex and type of occupation and comparison with 2014/15 Québec's population health survey (QPHS)

| | Total | Sex | | Type of occupation | | | |
|---|---|---|---|---|---|---|---|
| | N = 8220 | Men<br>n = 1375 | Women<br>n = 6845 | Nursing<br>n = 2140 | HCSW<br>n = 1572 | Physicians<br>n = 381 | Admin<br>n = 955 |
| Psychosocial risks | % | % | % | % | % | % | % |
| High psychological demands | 38.6 | 36.6 | 39.1 | 52.2 | 41.2 | 36.2 | 31.4 |
| QPHS 2014/15 all active workers | 32.2 | 30.1 | 34.5 | | | | |
| QPHS 2014/15 HCSA workers | 40.3 | 33.9 | 41.9 | | | | |
| Low or moderate decision authority | 52.1 | 49.2 | 52.7 | 57.5 | 61.6 | 34.9 | 37.2 |
| QPHS 2014/15 all active workers | 30.6 | 26.8 | 34.8 | | | | |
| QPHS 2014/15 HCSA workers | 36.0 | 31.9 | 37.0 | | | | |
| Low reward | 33.8 | 31.0 | 34.3 | 37.6 | 42.8 | 4.7 | 27.9 |
| QPHS 2014/15 all active workers | 20.9 | 18.9 | 23.1 | | | | |
| QPHS 2014/15 HCSA workers | 24.1 | 21.5 | 24.7 | | | | |

| | Total<br>N = 8220 | Sex | | Type of occupation | | | |
|---|---|---|---|---|---|---|---|
| | | Men<br>n = 1375 | Women<br>n = 6845 | Nursing<br>n = 2140 | HCSW<br>n = 1572 | Physicians<br>n = 381 | Admin<br>n = 955 |
| Low or moderate co-worker support | 17.2 | 15.9 | 17.4 | 13.7 | 23.0 | 8.4 | 18.5 |
| QPHS 2014/15 all active workers | 18.9 | 18.4 | 19.4 | | | | |
| QPHS 2014/15 HCSA workers | 18.1 | 17.8 | 18.1 | | | | |
| Low or moderate supervisor support | 22.5 | 21.0 | 22.9 | 27.0 | 24.7 | 12.6 | 17.1 |
| QPHS 2014/15 all active workers | 21.2 | 21.1 | 21.2 | | | | |
| QPHS 2014/15 HCSA workers | 23.9 | 22.9 | 24.1 | | | | |
| Difficult work-life balance | 30.8 | 28.2 | 31.3 | 40.7 | 23.2 | 47.5 | 24.8 |
| Not the means to do quality work | 77.5 | 74.6 | 78.1 | 87.2 | 69.2 | 79.5 | 73.9 |
| Work against their professional conscience | 52.2 | 48.0 | 53.1 | 66.9 | 53.4 | 52.0 | 36.9 |

Abbreviations: Adm= Administration and management staff; HCSW= healthcare support workers; HCSA workers= healthcare and social assistance workers; Nursing= nurses and nursing assistants; QPHS= Québec's population health survey

[a] Source : Institut national de santé publique du Québec. Risques psychosociaux du travail : définition, indicateurs et résultats de l'Enquête québécoise sur la santé de la population 2014-2015. Institut national de santé publique du Québec. 2021; en préparation.

**Table S4**. Prevalence ratios of high (Kessler scale score ≥ 7) work-related psychological distress according to the perception of the risk of acquiring COVID-19 at work and of some infection prevention and control measures

| Risks factors evaluated | Prevalence (proportion who agree) | | HIGH work-related psychological distress | |
|---|---|---|---|---|
| | Covid-19 HCWs | Non-COVID-19 HCWs | PR[a] | 95 % CI |
| High or very high perceived risk of acquiring COVID-19 at work (reference= low or moderate) | 29.2% | 15.7% | 1.3 | 1.2 – 1.4 |
| Personal protective equipment not available | 4.7% | 3.6% | 0.9 | 0.8 – 1.1 |
| Insufficient infection prevention and control training | 11.8% | 13.5% | 1.1 | 1.0 – 1.2 |
| Insufficient practical training about the use of personal protective equipment | 26.6% | 35.7% | 1.0 | 1.0 – 1.1 |
| Lack of information about identification of COVID-19 confirmed or suspected patients | 17.5% | 18.4% | 1.2 | 1.1 – 1.2 |
| Difficult access to COVID-19 screening test | 9.0% | 7.2% | 1.1 | 1.0 – 1.2 |
| Physical environment not allowing physical distancing during breaks and meals | 19.6% | 22.4% | 1.2 | 1.1 – 1.3 |
| Insufficient human resources to ensure patient care and HCW safety | 36.4% | 39.5% | 1.5 | 1.4 – 1.6 |
| Persistent COVID-19 symptoms longer than 4 weeks | 49.2% | NA | 1.3 | 1.2 – 1.4 |
| COVID-19 status (reference= non-COVID-19) | 51.8% | 48.2 | 0.7 | 0.7 – 0.8 |

Abbreviations: CI= confidence interval; HCW=Healthcare worker; PR= Prevalence ratio;

[a] A robust Poisson model including all risk factors and adjusted for sex, age (18-44, 45-59, ≥60 years), race/ethnicity (white, black, other) and type of occupation (nursing, healthcare support worker, physician, administration and management staff, other). An interaction strategy was used to account for structural missing values for persistent COVID-19 symptoms among non-COVID-19 healthcare workers.
